# No Overall Survival Benefit with Adding Chemotherapy to Immunotherapy in PD-L1 TPS ≥ 50% NSCLC: An Agent-Stratified Reassessment

**DOI:** 10.64898/2026.09.01.26361919

**Authors:** Fujun Han, Jinwen Wang, Songchen Shi, Meile Jin, Chunhui Ren

## Abstract

**IMPORTANCE:** A recent meta-analysis showed that chemoimmunotherapy was associated with improved overall survival (OS) compared with immune checkpoint inhibitor (ICI) monotherapy for programmed death-ligand 1 (PD-L1) tumor proportion score (TPS) ≥ 50% advanced non-small-cell lung cancer (NSCLC). However, whether this benefit reflects chemotherapy effect or ICI heterogeneity remains unclear.

**OBJECTIVE:** To reassess the survival benefit of adding chemotherapy to ICI monotherapy using agent-stratified comparisons anchored to chemotherapy.

**DATA SOURCES:** The 24 phase 3 randomized clinical trials included in the original meta-analysis (search date, August 3, 2025).

**DATA EXTRACTION AND SYNTHESIS:** Hazard ratios (HRs) for OS and progression-free survival (PFS) were extracted from each trial in the original meta-analysis. Two analytic frameworks were used: within-agent comparisons (same ICI in both chemoimmunotherapy and monotherapy) and across-agent comparisons (ICI in one treatment strategy only). For within-agent comparisons, a two-stage random-effects meta-analysis was conducted. In stage 1, ICI-specific HRs for chemoimmunotherapy and ICI monotherapy versus chemotherapy were pooled and their ratio was calculated (RHR = HRchemoimmuno/HRmono; RHR < 1 favors chemoimmunotherapy). The RHRs were pooled in stage 2. For across-agent comparisons, RHR was derived from pooled HRs by treatment strategy.

**MAIN OUTCOMES AND MEASURES:** Endpoints were OS and PFS.

**RESULTS:** In within-agent comparisons (4 ICIs; 13 trials; N = 3252), pooled RHR was 0.94 (95% CI, 0.78–1.13; *P* = .48; *I*² = 0.0%) for OS and 0.85 (95% CI, 0.68–1.06; *P* = .14; *I²* = 0.0%) for PFS. In across-agent comparisons (7 ICIs; 11 trials; N = 2231), RHR favored chemoimmunotherapy for OS (0.68; 95% CI, 0.50–0.92; *P* = .01) and PFS (0.46; 95% CI, 0.37–0.58; *P* < .001). In a sensitivity analysis restricted to trials of NCCN-recommended regimens, pooled RHR was 1.02 (95% CI, 0.81–1.28; *P* = .87) for OS.

**CONCLUSIONS AND RELEVANCE:** In the within-agent comparisons, adding chemotherapy to ICI monotherapy did not improve OS or PFS in patients with PD-L1 TPS ≥ 50% advanced NSCLC. The benefit in the original meta-analysis appears driven by across-ICI heterogeneity. These findings are consistent with ICI monotherapy as a standard first-line option and underscore the need for agent-level stratification in across-trial comparisons.

## Introduction

For patients with advanced non-small cell lung cancer (NSCLC) and programmed death-ligand 1 (PD-L1) tumor proportion score (TPS) of 50% or higher, the National Comprehensive Cancer Network (NCCN) recommends both immune checkpoint inhibitor (ICI) monotherapy (eg, pembrolizumab, cemiplimab) and combination with chemotherapy as Category 1 options.^1^ In the recent meta-analysis by Di Federico and colleagues, chemoimmunotherapy was associated with significantly improved overall survival (OS) and progression-free survival (PFS) compared with ICI monotherapy,² providing comprehensive evidence informing first-line treatment selection in this population.

The original report used two complementary analyses,^2^ both of which are sensitive to potential confounding from ICI-specific and study-level heterogeneity. First, the meta-analysis pooled data across different ICIs without stratifying by ICI agent. If treatment effects varied across ICIs and these ICIs were unevenly distributed across trials evaluating monotherapy and chemoimmunotherapy, the pooled estimate could be confounded by across-ICI differences rather than reflect the incremental effect of adding chemotherapy. Second, a reconstructed individual patient data (IPD) analysis compared patients directly across trials without anchoring the comparison to a common chemotherapy control. Such unanchored cross-trial comparisons may not reliably distinguish the incremental effect of adding chemotherapy from systematic differences in trial populations, clinical settings, subsequent therapies, and other study-level factors, and may therefore yield estimates influenced by between-trial heterogeneity.

To address these concerns, we adopted a two-stage meta-analytic framework for indirect comparison, stratified by individual ICI agent and anchored to the common chemotherapy comparator. In stage 1, we separately pooled the hazard ratios (HRs) for chemoimmunotherapy versus chemotherapy and ICI monotherapy versus chemotherapy within each ICI, and then calculated the ratio of these HRs (RHR), an established metric for indirect treatment comparisons anchored to a common control.^3,4^ In stage 2, these ICI-specific RHRs were pooled to generate a summary estimate. This approach tests whether the reported survival benefit is better explained by chemotherapy or by ICI heterogeneity.

## Methods

This was a secondary meta-analysis designed to reassess the comparative efficacy of chemoimmunotherapy versus ICI monotherapy in patients with PD-L1 TPS ≥ 50% advanced NSCLC. It is reported in accordance with the applicable items of the Preferred Reporting Items for Systematic Reviews and Meta-Analyses (PRISMA) 2020 statement.^5^

### Data Source

We used the same set of 24 phase 3 randomized trials included in the original meta-analysis by Di Federico and colleagues,^2^ which searched PubMed, Embase, and major oncology conference proceedings for studies published before August 3, 2025. The inclusion criteria were: (1) phase 3 randomized trials; (2) inclusion of patients with previously untreated advanced NSCLC; (3) comparison of a PD-L1 inhibitor, either as monotherapy or in combination with platinum-based chemotherapy, vs platinum-based chemotherapy alone; (4) reporting of HRs and 95% confidence intervals (CIs) for patients with high PD-L1 expression (PD-L1TPS ≥ 50%).

HRs for overall survival (OS) and progression-free survival (PFS) were extracted from the original meta-analysis. Additional trial-level characteristics were extracted using a standardized form, independently by two reviewers, and discrepancies were reconciled by consensus or referral to a senior author. No individual patient data were reconstructed.

## Statistical Analysis

### Meta-analysis

Given the anticipated clinical and methodological heterogeneity across trials and individual ICIs, meta-analyses used inverse-variance weighting with a random-effects model. The between-study variance, τ^2^, was estimated using restricted maximum likelihood. Statistical heterogeneity was quantified using the *I*² statistic and assessed using the Cochran Q test. To replicate the original meta-analysis, both fixed-effects and random-effects models were applied.

### Ratio of Hazard Ratios (RHR)

To compare the treatment effects of chemoimmunotherapy versus monotherapy within the same ICI agent, we calculated RHR, defined as HRchemoimmuno/HRmono (RHR < 1 favors chemoimmunotherapy), where both HRs are estimated versus the chemotherapy comparator. For each ICI agent, the agent-specific HRs were obtained by pooling eligible trials; if only one trial was available, the reported HR was used directly. This comparison followed the framework described by Bucher et al,^3^ with variance calculations based on Altman and Bland’s method for comparing two independent estimates.^4^ RHRs were calculated on the log scale, and 95% CIs were derived using the delta method under a normal approximation. A two-sided *P* value was obtained from a z test of the log-transformed ratio. Statistically, testing this RHR against the null is equivalent to a test for subgroup differences in a meta-analysis.^6^

### Analytical Framework

Trials were classified into two analytical frameworks based on whether the same ICI was evaluated in both chemoimmunotherapy and ICI monotherapy (Figure 1). Within-agent comparisons: For ICIs with trials in both treatment strategies (pembrolizumab, atezolizumab, cemiplimab, and durvalumab), we conducted chemotherapy-anchored indirect comparisons via a two-stage meta-analysis. In stage 1, within each agent, HRs for the two strategies versus chemotherapy were pooled separately; if only one trial was available, the reported HR was used directly. The RHR was then calculated. In stage 2, the agent-specific RHRs were pooled to generate a summary within-agent estimate. This approach estimates the treatment effect within each agent while minimizing the potential confounding present in the unstratified approach. Across-agent comparisons: For ICIs with data available in only one treatment strategy, trials were pooled within each strategy to derive overall HRs, from which an across-agent RHR was calculated.

**Figure 1.**
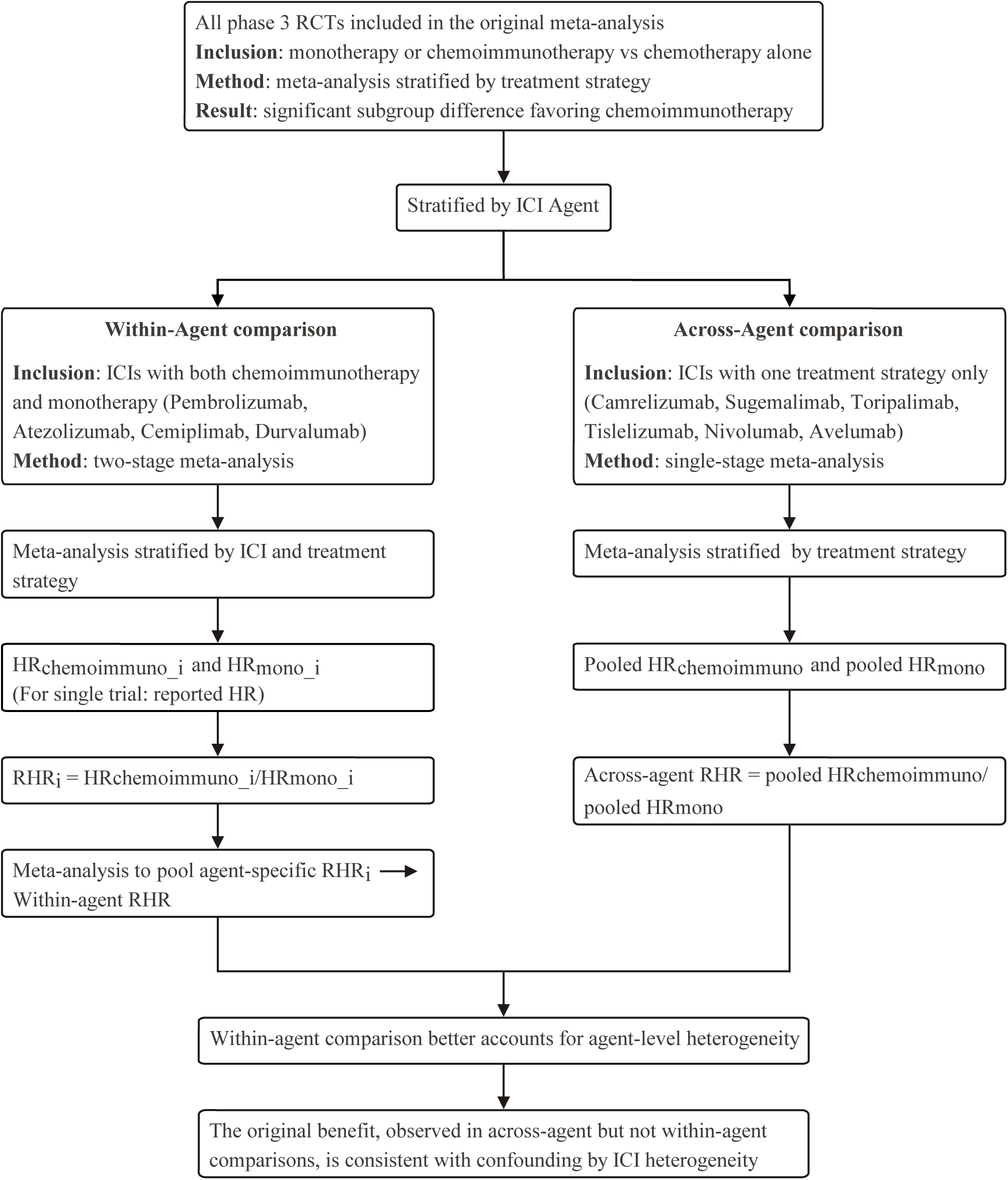
Analytic Framework for Within-Agent and Across-Agent Comparisons. Abbreviations: HR, hazard ratio; ICI, immune checkpoint inhibitor; RHR, ratio of hazard ratios (HRchemoimmuno/HRmono; RHR < 1 favors chemoimmunotherapy).

### Overall Analysis for Replication of the Original Meta-Analysis

To establish comparability with the original meta-analysis, we performed meta-analyses using both fixed-effects and random-effects models to obtain pooled HRs for chemoimmunotherapy versus chemotherapy and for ICI monotherapy versus chemotherapy across all trials, regardless of ICI agent. The fixed-effect model was the primary analytic approach used in the original report. We also conducted the Cochran Q test for subgroup differences between the two treatment-strategy estimates, as in the original meta-analysis. These analyses were designed to reproduce the original meta-analysis and served as the reference. Our agent-stratified findings were compared against this reference to evaluate whether the aggregate effect was confounded by ICI-specific heterogeneity.

### Sensitivity Analysis

To ensure clinical relevance, we performed a sensitivity analysis restricted to trials of the current NCCN-recommended first-line regimens for PD-L1 TPS ≥ 50% advanced NSCLC.^1^ A leave-one-out sensitivity analysis was also conducted to assess the robustness of the findings.

All statistical tests were two-sided, with *P* < .05 considered statistically significant. Analyses were performed using R, version 4.4.2, with the ‘meta’ package.

## Results

### Trial Characteristics

A total of 24 phase 3 randomized clinical trials comprising 5483 patients from the original meta-analysis were included (Table 1)^2,7–30^: 16 evaluated chemoimmunotherapy and 8 evaluated ICI monotherapy. 4 ICIs were represented in both treatment strategies, comprising 13 trials: pembrolizumab (2 monotherapy and 2 chemoimmunotherapy trials), atezolizumab (1 and 4), cemiplimab (1 and 1), and durvalumab (1 and 1). All of these trials were global. Among the 5 monotherapy trials, 4 were positive and 1 was negative; among the 8 chemoimmunotherapy trials, 4 were positive and 4 were negative. 7 ICIs were represented in only one strategy, comprising 11 trials: 3 monotherapy trials (2 nivolumab, 1 avelumab) were global and negative; 8 chemoimmunotherapy trials (camrelizumab, toripalimab, sugemalimab, sintilimab, tislelizumab) were China only and all positive.

**Table 1.**
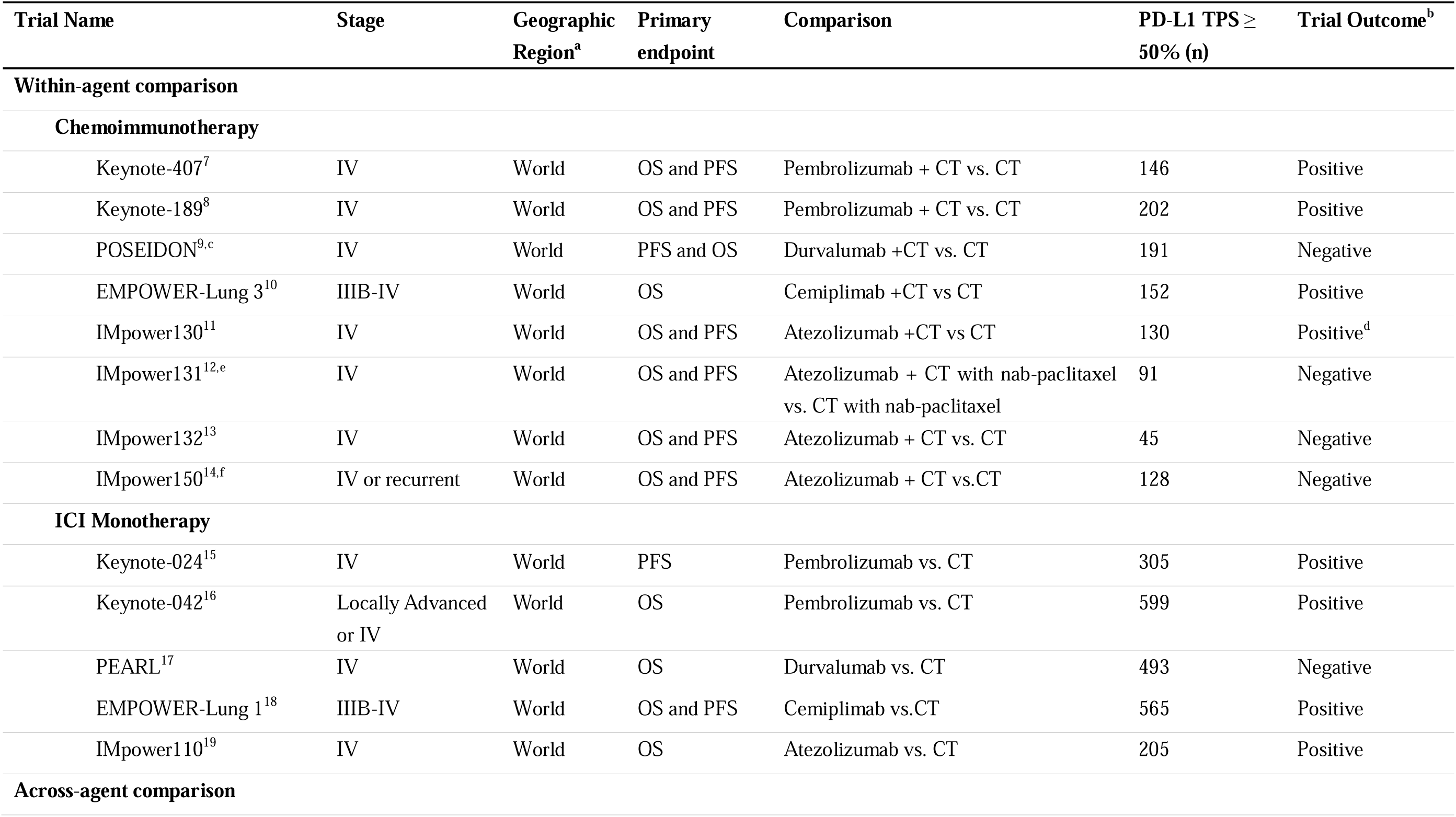
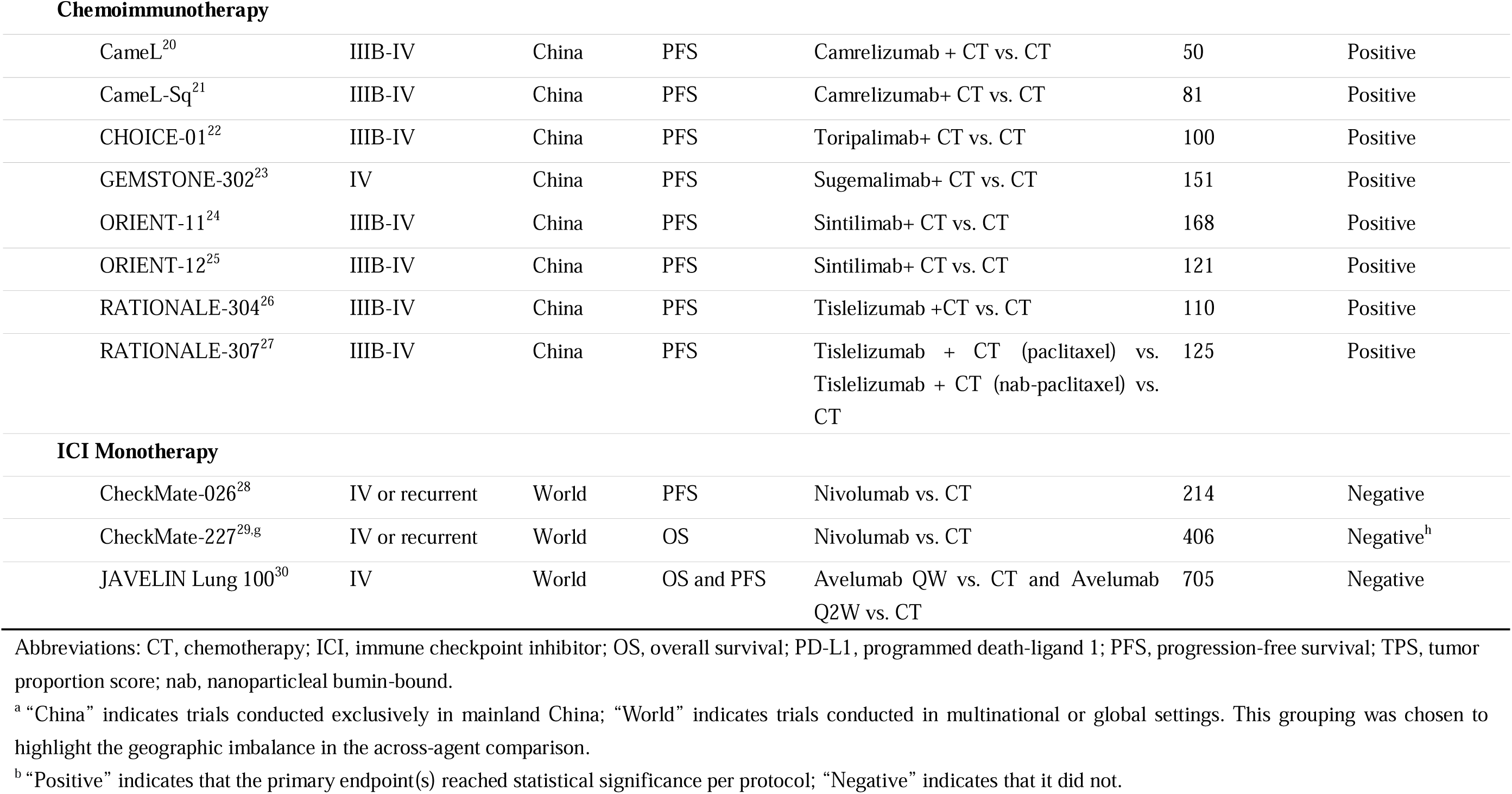
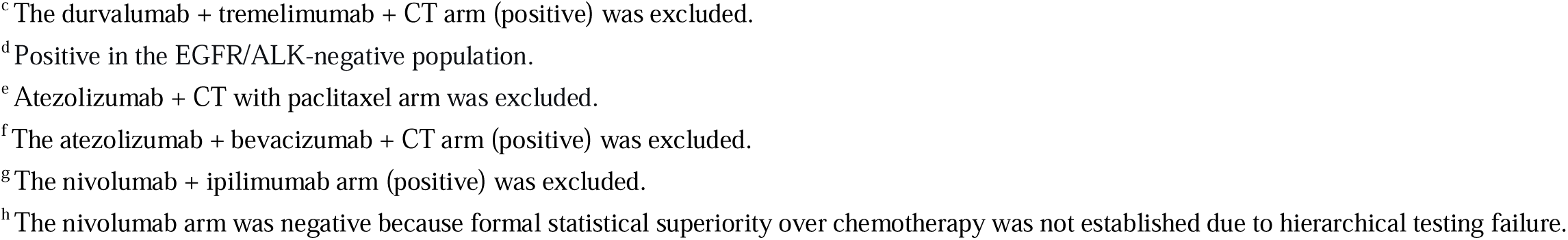
Baseline Characteristics of Included Trials.

### Overall Analysis

To replicate the original meta-analysis, we performed an overall meta-analysis unstratified by ICI agent, and the findings were consistent with the original report (eTables 1-4 in the Supplement). For OS, the pooled HR was 0.63 (95% CI, 0.56–0.72) for chemoimmunotherapy and 0.74 (95% CI, 0.67–0.82) for ICI monotherapy under the fixed-effect model (the primary analytic approach used in the original meta-analysis). The test for subgroup differences yielded χ² = 4.13 (*P* = .04; *I*² = 75.8%) under the fixed-effect model and Q = 3.80 (*P* = .05; *I*² = 73.7%) under the random-effects model.

To provide a unified metric for this comparison, we derived the overall RHR from these pooled estimates (Figures 2 and 3). For OS, the overall RHR was 0.85 (95% CI, 0.73–1.00; *P* = .05), where both component HRs were pooled under the random-effects model. This RHR-based test was consistent with the subgroup difference test under the same model (Q = 3.80; *P* = .05).

**Figure 2.**
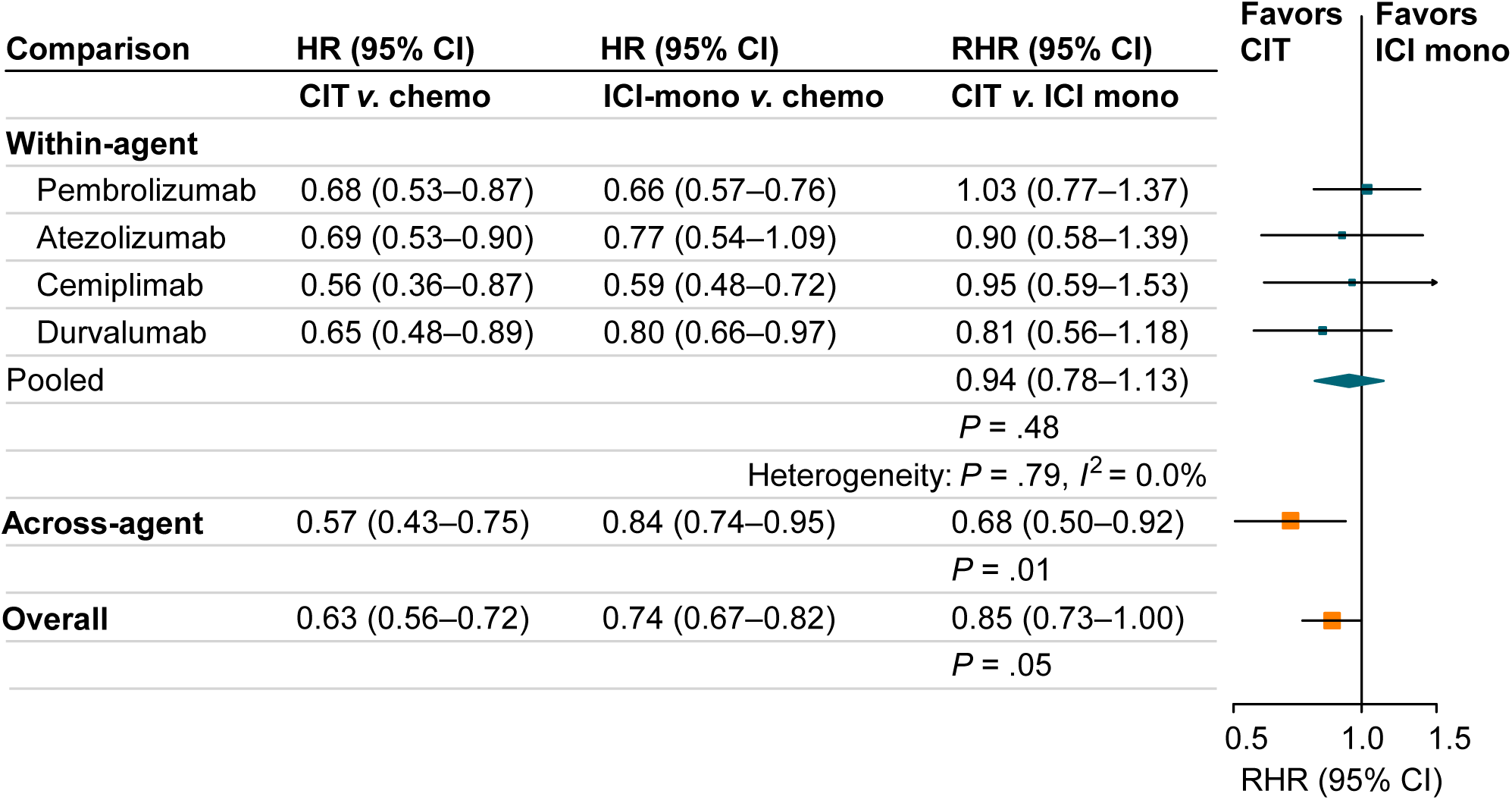
Overall Survival for Within-Agent vs Across-Agent Comparisons. Abbreviations: CIT, chemoimmunotherapy; HR, hazard ratio; ICI, immune checkpoint inhibitor. Forest plots of individual trial HRs and pooled ICI-specific HRs for chemoimmunotherapy vs chemotherapy and for ICI monotherapy vs chemotherapy are shown in eFigures 1 and 2. The ratio of HRs (RHR) comparing chemoimmunotherapy with monotherapy was calculated as HRchemoimmuno/HRmono; RHR < 1 favors chemoimmunotherapy. For within-agent comparisons (ICIs with trials in both treatment strategies), a two-stage approach was used: agent-specific HRs were first pooled by treatment strategy within each ICI; agent-specific RHRs were then pooled to obtain the summary estimate. For across-agent comparisons (ICIs with trials in only one strategy) and for the overall comparison (all trials regardless of ICI agent), HRs were pooled directly by treatment strategy, and a single RHR was derived from these pooled estimates.

**Figure 3.**
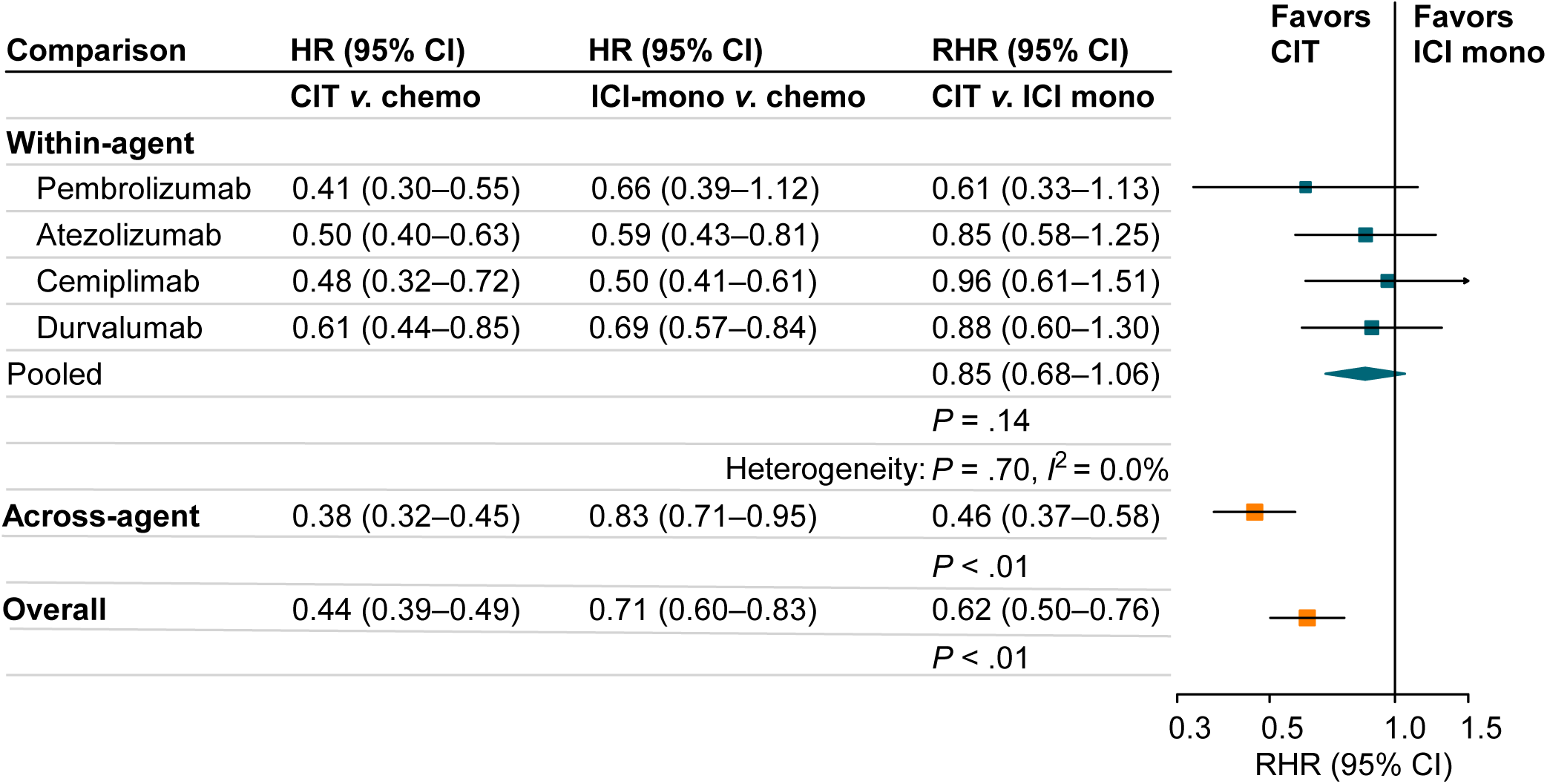
Progression-Free Survival for Within-Agent vs Across-Agent Comparisons. Abbreviations: CIT, chemoimmunotherapy; HR, hazard ratio; ICI, immune checkpoint inhibitor. Forest plots of individual trial HRs and pooled ICI-specific HRs for chemoimmunotherapy vs chemotherapy and for ICI monotherapy vs chemotherapy are shown in eFigures 3 and 4. The ratio of HRs (RHR) comparing chemoimmunotherapy with monotherapy was calculated as HRchemoimmuno/HRmono; RHR < 1 favors chemoimmunotherapy. For within-agent comparisons (ICIs with trials in both treatment strategies), a two-stage approach was used: agent-specific HRs were first pooled by treatment strategy within each ICI; agent-specific RHRs were then pooled to obtain the summary estimate. For across-agent comparisons (ICIs with trials in only one strategy) and for the overall comparison (all trials regardless of ICI agent), HRs were pooled directly by treatment strategy, and a single RHR was derived from these pooled estimates.

### Within-Agent Comparisons

Among the 4 ICIs with data for both treatment strategies (13 trials; 3252 patients), individual ICI-specific RHRs for OS ranged from 0.81 to 1.03, with all 95% CIs crossing 1.0 (Figure 2). The pooled RHR was 0.94 (95% CI, 0.78–1.13; *P* = .48; *I*² = 0.0%). For PFS, individual ICI-specific RHR estimates also had 95% CIs crossing 1.0 (Figure 3), and the pooled RHR was 0.85 (95% CI, 0.68–1.06; *P* = .14; *I*² = 0.0%).

### Across-agent comparisons

In across-agent comparisons, which included the remaining 7 ICIs with 11 trials (2231 patients) evaluating ICIs with only one treatment strategy, chemoimmunotherapy was associated with a significant benefit for both OS (RHR = 0.68; 95% CI, 0.50–0.92; *P* = .01) and PFS (RHR = 0.46; 95% CI, 0.37–0.58; *P* < .001).

### Sensitivity Analyses

To assess the robustness of the within-agent findings, we performed two sensitivity analyses (Figure 4). When restricted to NCCN-recommended first-line regimens for PD-L1 TPS ≥ 50% advanced NSCLC (Version 6.2026)—pembrolizumab, atezolizumab, and cemiplimab—the pooled RHR was 1.02 (95% CI, 0.81–1.28; *P* = .87) for OS and 0.83 (95% CI, 0.62–1.12; *P* = .23) for PFS. Leave-one-out sensitivity analyses yielded RHRs ranging from 0.87 to 0.98 for OS and 0.82 to 0.89 for PFS, with all 95% CIs crossing 1.0.

**Figure 4.**
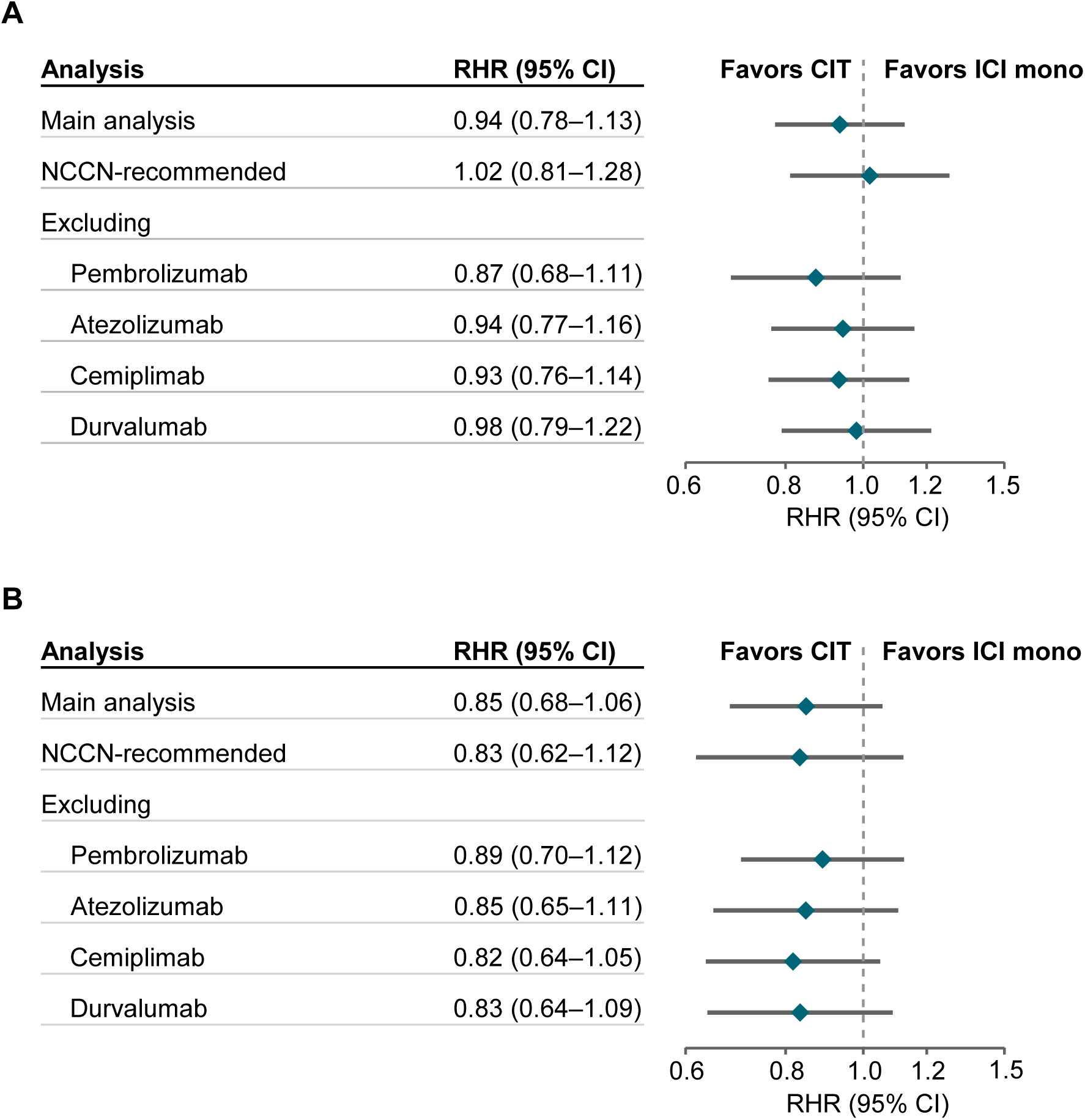
Sensitivity Analyses of NCCN-Recommended Regimens and Leave-One-Out for Within-Agent Findings. Forest plot for overall survival (A) and progression-free survival (B). Abbreviations: ICI, immune checkpoint inhibitor; RHR, ratio of hazard ratios (HRchemoimmuno/HRmono; RHR < 1 favors chemoimmunotherapy). Trials underlying NCCN-recommended regimens (Version 6.2026) included the three within-agent ICIs: pembrolizumab (KEYNOTE-024, −042, −189, −407), cemiplimab (EMPOWER-Lung 1 and −3), and atezolizumab (IMpower110 and −130).

## Discussion

In this secondary meta-analysis of 24 randomized trials including 5483 patients with PD-L1 TPS ≥ 50% NSCLC, within-agent comparisons, in which the same ICI was evaluated in both monotherapy and chemoimmunotherapy, showed no significant OS or PFS benefit with adding chemotherapy. In contrast, across-agent comparisons, which included ICIs available in only one treatment strategy, showed a significant benefit favoring chemoimmunotherapy for both endpoints. The apparent benefit observed in the overall analysis, which mirrored the original meta-analysis conclusions, was not reproduced in the within-agent comparisons but persisted in the across-agent comparisons.

The two analytic frameworks differ in their susceptibility to confounding. Within-agent comparisons hold the ICI agent constant, whereas across-agent comparisons do not. The within-agent approach also benefits from greater homogeneity within trials of the same agent series (eg, KEYNOTE),^7,8,15,16^ which share similar designs and used the same PD-L1 testing method (the PD-L1 IHC 22C3 pharmDx assay). The across-agent comparison estimates the average difference between two groups of trials that evaluated different ICIs and is therefore susceptible to confounding by ICI-specific effects. In the across-agent analysis, chemoimmunotherapy trials, including CameL, CameL-Sq, CHOICE-01, GEMSTONE-302, and RATIONALE-304, reported HRs for OS ranging from 0.38 to 0.82 (pooled HR, 0.57; 95% CI, 0.43–0.75, eFigure 1). In contrast, monotherapy trials, including CheckMate 026, CheckMate 227, and JAVELIN Lung 100, reported HRs for OS ranging from 0.82 to 0.90 (pooled HR, 0.84; 95% CI, 0.74–0.95, eFigure 2). Notably, all monotherapy trials in the across-agent comparison were global, whereas the chemoimmunotherapy trials were predominantly China-only (Table1). The stark contrast between the two pooled estimates underscores the difficulty of determining whether the apparent benefit in the across-agent comparison reflects differences between the two sets of trials or a genuine chemotherapy effect. Therefore, this pattern, overall benefit observed in across-agent but not within-agent comparisons, is consistent with confounding by ICI- and trial-level heterogeneity rather than a true chemotherapy effect.

Our analysis provides clinically relevant evidence that ICI monotherapy remains a reasonable standard of care. In the within-agent comparisons, a non-significant trend toward PFS benefit did not translate into an OS gain. The pooled OS RHR was 0.94 (95% CI, 0.78–1.13). While a larger sample might detect a small OS benefit, the magnitude would likely fall below clinically meaningful benefit, particularly when weighed against the well-established toxicities of chemotherapy. When restricted to trials of NCCN-recommended first-line regimens (pembrolizumab, atezolizumab, and cemiplimab),^1^ the pooled OS RHR was 1.02 (95% CI, 0.81–1.28), further supporting the absence of a meaningful OS advantage with the addition of chemotherapy.

Several limitations should be acknowledged. First, while a PFS trend favored chemoimmunotherapy without an OS gain, this dissociation may reflect a transient effect of chemotherapy that attenuates over time. Formal testing of this temporal dynamic (eg, via landmark or restricted mean survival time analyses) would require individual patient data reconstruction, which was beyond the scope of this aggregate-level methodological reassessment but represents a valuable direction for further validation. Second, the distinction between within-agent and across-agent comparisons should not be overinterpreted as evidence of differential efficacy across ICI agents. The within-agent comparisons should be understood as within-series comparisons: for each ICI, the monotherapy and chemoimmunotherapy trials belong to the same trial program (eg, KEYNOTE series), sharing comparatively consistent PD-L1 assays, eligibility criteria, and assessment procedures. The RHR metric, by taking the ratio of the two HRs, effectively minimizes these shared trial-level factors, isolating the incremental effect of adding chemotherapy. Thus, our findings illustrate confounding by across-series heterogeneity in the unstratified analysis, rather than a comparative assessment of individual ICIs. Third, although the within-agent approach minimized confounding across trials by restricting comparisons to the same agent, this remains an indirect comparison based on trial-level data, limiting adjustment for patient-level covariates. Additionally, the absence of statistical heterogeneity, as measured by *I*², does not preclude clinically meaningful differences across trials in patient populations, study designs, or treatment protocols. Despite these limitations, our findings provide an important methodological perspective on the interpretation of between-trial comparisons.

## Conclusion

In this secondary meta-analysis restricted to within-agent comparisons, adding chemotherapy to ICI monotherapy was not associated with a statistically significant OS or PFS benefit in patients with PD-L1 TPS ≥ 50% advanced NSCLC. The apparent benefit observed in the original meta-analysis was likely driven by across-agent heterogeneity rather than a true chemotherapy effect. These findings are consistent with ICI monotherapy as a standard first-line option to avoid chemotherapy-related toxicity and underscore the critical importance of agent-level stratification in indirect treatment comparisons.

## Data Availability

All data produced in the present study are available upon reasonable request to the authors

## Acknowledgment

The authors thank Ph.D. Suli Liu (College of Mathematics, Jilin University) for her expert statistical guidance.

## Author Contributions

Drs Han and Wang had full access to all of the data in the study and take responsibility for the integrity of the data and the accuracy of the data analysis. Concept and design: Fujun Han. Acquisition, analysis, or interpretation of data: All authors. Drafting of the manuscript: Fujun Han, Jinwen Wang. Critical review of the manuscript for important intellectual content: All authors. Statistical analysis: Jinwen Wang, Meile Jin, Songchen Shi, Fujun Han. Obtained funding: Fujun Han. Administrative, technical, or material support: Fujun Han. Supervision: Fujun Han.

## Data Sharing Statement

The scripts and extracted data (hazard ratios and 95% confidence intervals) are available on reasonable request to the corresponding author.

