## Supplement for "No Overall Survival Benefit with Adding Chemotherapy to Immunotherapy in PD-L1 TPS ≥ 50% NSCLC: An Agent-Stratified Reassessment"

**eTable 1.** Replication of Original Meta-Analysis Results for Overall Survival

**eTable 2.** Replication of Subgroup Difference Test Between Chemoimmunotherapy and ICI Monotherapy by Statistical Model for Overall Survival

**eTable 3.** Replication of Original Meta-Analysis Results for Progression-Free Survival
**eTable 4.** Replication of Subgroup Difference Test Between Chemoimmunotherapy and ICI Monotherapy by Statistical Model for Progression-Free Survival

**eFigure 1.** Forest Plots of Hazard Ratios for Overall Survival Comparing Chemoimmunotherapy with Chemotherapy

**eFigure 2.** Forest Plots of Hazard Ratios for Overall Survival Comparing ICI Monotherapy with Chemotherapy

**eFigure 3.** Forest Plots of Hazard Ratios for Progression-Free Survival Comparing Chemoimmunotherapy with Chemotherapy

**eFigure 4.** Forest Plots of Hazard Ratios for Progression-Free Survival Comparing ICI Monotherapy with Chemotherapy

**eTable 1.** Replication of Original Meta-Analysis Results for Overall Survival

| **Regimen** | **Source** | **HR** | **95% CI** | **Heterogeneity** |
| --- | --- | --- | --- | --- |
| **Fixed-effects** | | | | |
| Chemoimmunotherapy | Original | 0.63 | 0.56–0.72 | χ² = 8.59; *I*² = 0.0% |
|  | Replication | 0.63 | 0.56–0.72 | χ² = 8.97; *I*² = 0.0% |
| ICI Monotherapy | Original | 0.74 | 0.69–0.80 | χ² = 12.47; *I*² = 36.0% |
|  | Replication | 0.74 | 0.69–0.80 | χ² = 12.41; *I*² = 35.5% |
| **Random-effects** | | | | |
| Chemoimmunotherapy | Original | 0.63 | 0.56–0.72 | χ² = 8.59; *I*² = 0.0% |
|  | Replication | 0.63 | 0.56–0.72 | χ² = 8.97; *I*² = 0.0% |
| ICI Monotherapy | Original | 0.74 | 0.68–0.82 | χ² = 12.47; *I*² = 36.0% |
|  | Replication | 0.74 | 0.67–0.82 | χ² = 12.41; *I*² = 35.5% |

Abbreviations: CI, confidence interval; HR, hazard ratio; *I*², measure of heterogeneity; χ², Cochran’s Q statistic (reported as χ² for fixed-effect model).

Note: All estimates are presented to two decimal places for HRs and CIs, and to one decimal place for *I*² values, consistent with the original reporting format. The original meta-analysis was performed using Review Manager, version 5.4 (Cochrane). Our replication was performed using R, version 4.4.2, with the ‘meta’ package. Minor discrepancies in χ² statistics between original and replicated analyses might be attributable to rounding and differences in estimation algorithms between the two software platforms. All point estimates and statistical inferences remain consistent.

**eTable 2.** Replication of Subgroup Difference Test Between Chemoimmunotherapy and ICI Monotherapy by Statistical Model for Overall Survival

| **Statistical Model** | **Source** | **Test Statistic (χ²/Q)** | ***P* value** | ***I*²** |
| --- | --- | --- | --- | --- |
| Fixed-effects | Original | 4.1 | .04 | 75.8% |
|  | Replication | 4.1 | .04 | 75.8% |
| Random-effects | Original | 3.82 | .05 | 73.8% |
|  | Replication | 3.80 | .05 | 73.7% |

Abbreviations: *I*², measure of heterogeneity; OS, overall survival; χ², Cochran’s Q statistic (reported as χ² for fixed-effect model).

Note: Rounding to one or two decimal places follows the original reporting format. All point estimates and statistical inferences remain consistent. See eTable 1 footnotes for other details.

**eTable 3.** Replication of Original Meta-Analysis Results for Progression-Free Survival

| **Regimen** | **Source** | **HR** | **95% CI** | **Heterogeneity** |
| --- | --- | --- | --- | --- |
| **Fixed-effects** | | | | |
| Chemoimmunotherapy | Original | 0.44 | 0.39–0.49 | χ² = 18.01; *I*² = 11% |
|  | Replication | 0.44 | 0.39–0.49 | χ² = 17.93; *I*² = 10.8% |
| ICI Monotherapy | Original | 0.70 | 0.65–0.76 | χ² = 33.65; *I*² = 76.0% |
|  | Replication | 0.70 | 0.65–0.76 | χ² = 34.21; *I*² = 76.6% |
| **Random-effects** | | | | |
| Chemoimmunotherapy | Original | 0.44 | 0.39–0.49 | χ² = 18.01; *I*² = 11.0% |
|  | Replication | 0.44 | 0.39–0.49 | χ² = 17.93; *I*² = 10.8% |
| ICI Monotherapy | Original | 0.71 | 0.60–0.83 | χ² = 33.65; *I*² = 76.0% |
|  | Replication | 0.71 | 0.60–0.83 | χ² = 34.21; *I*² = 76.6% |

Abbreviations: CI, confidence interval; HR, hazard ratio; *I*², measure of heterogeneity; χ², Cochran’s Q statistic (reported as χ² for fixed-effect model).

Note: See eTable 1 footnotes.

**eTable 4.** Replication of Subgroup Difference Test Between Chemoimmunotherapy and ICI Monotherapy by Statistical Model for Progression-Free Survival

| **Statistical Model** | **Source** | **Test Statistic (χ²/Q)** | ***P* value** | ***I*²** |
| --- | --- | --- | --- | --- |
| Fixed-effects | Original | 48.1 | < .001 | 97.9% |
|  | Replication | 48.3 | < .001 | 97.9% |
| Random-effects | Original | 23.46 | < .001 | 95.5% |
|  | Replication | 21.60 | < .001 | 95.4% |

Abbreviations: *I*², measure of heterogeneity; χ², Cochran’s Q statistic (reported as χ² for fixed-effect model).

Note: See eTable 2 footnotes.

**eFigure 1.** Forest Plots of Hazard Ratios for Overall Survival Comparing Chemoimmunotherapy with Chemotherapy
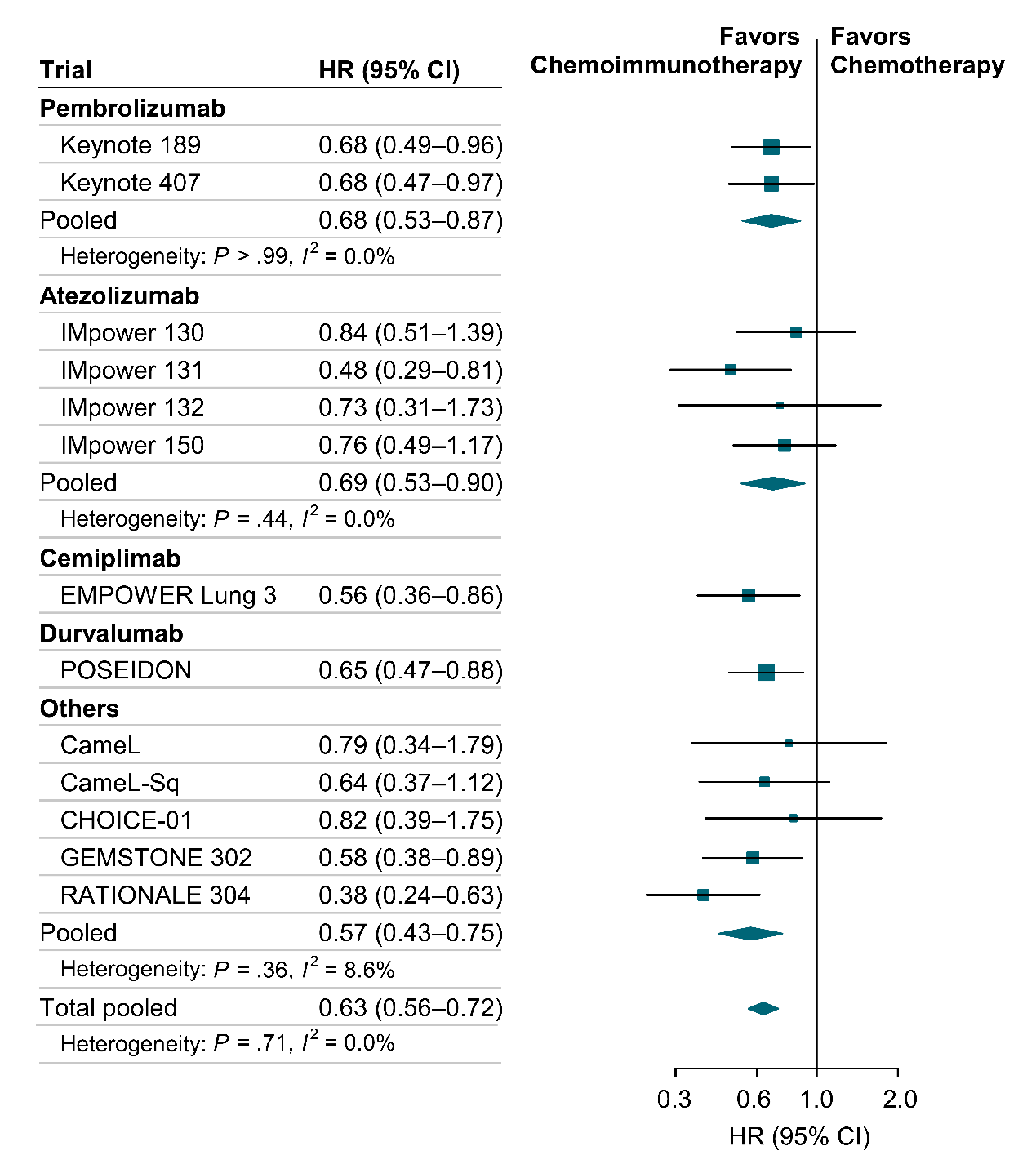

Abbreviations: CI, confidence interval; HR, hazard ratio; *I*², I-squared statistic (percentage of total variability due to heterogeneity).

**eFigure 2.** Forest Plots of Hazard Ratios for Overall Survival Comparing ICI Monotherapy with Chemotherapy
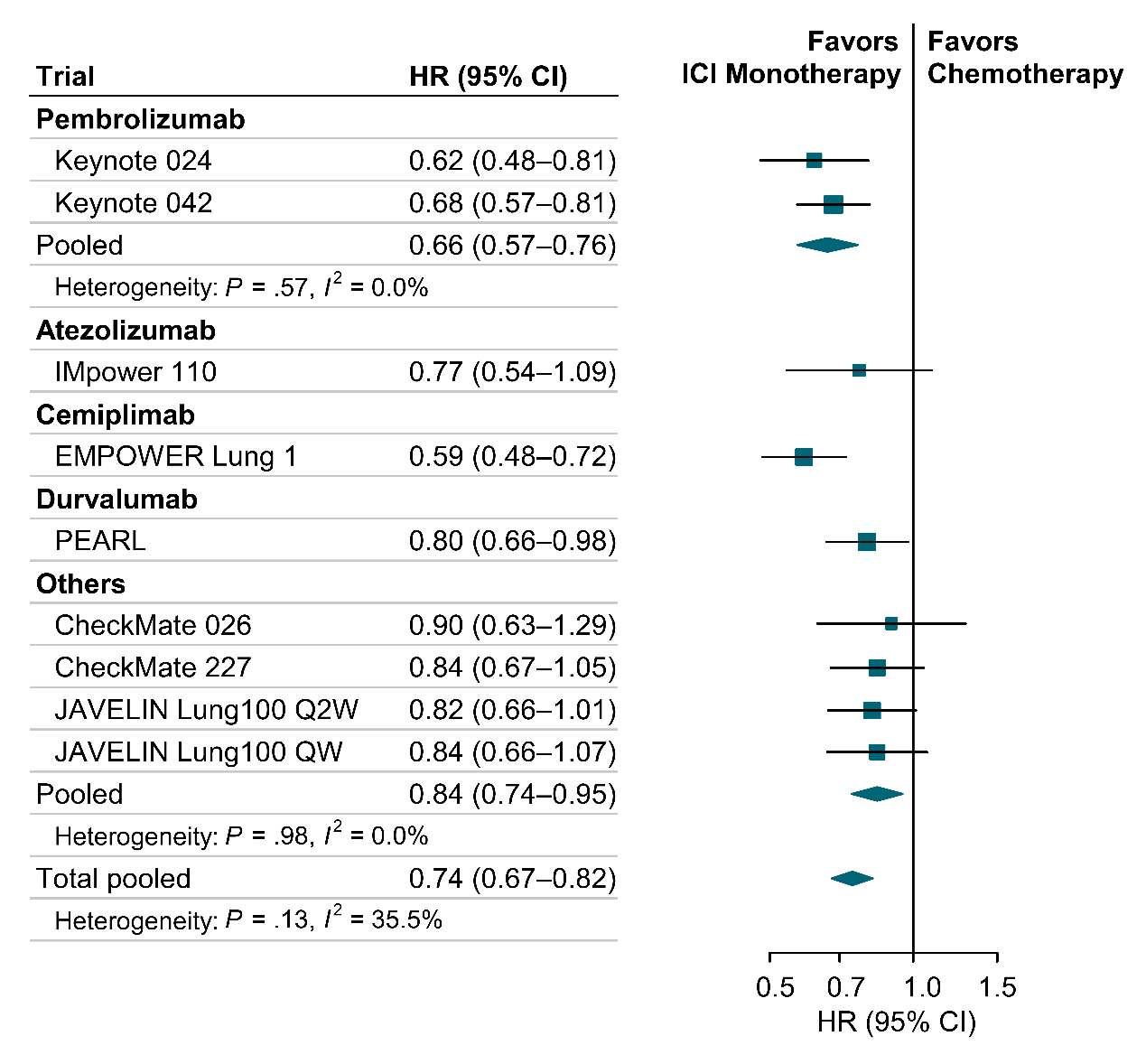
 Abbreviations: CI, confidence interval; HR, hazard ratio; ICI, immune checkpoint inhibitor; *I*², I-squared statistic (percentage of total variability due to heterogeneity).

**eFigure 3.** Forest Plots of Hazard Ratios for Progression-Free Survival Comparing Chemoimmunotherapy with Chemotherapy
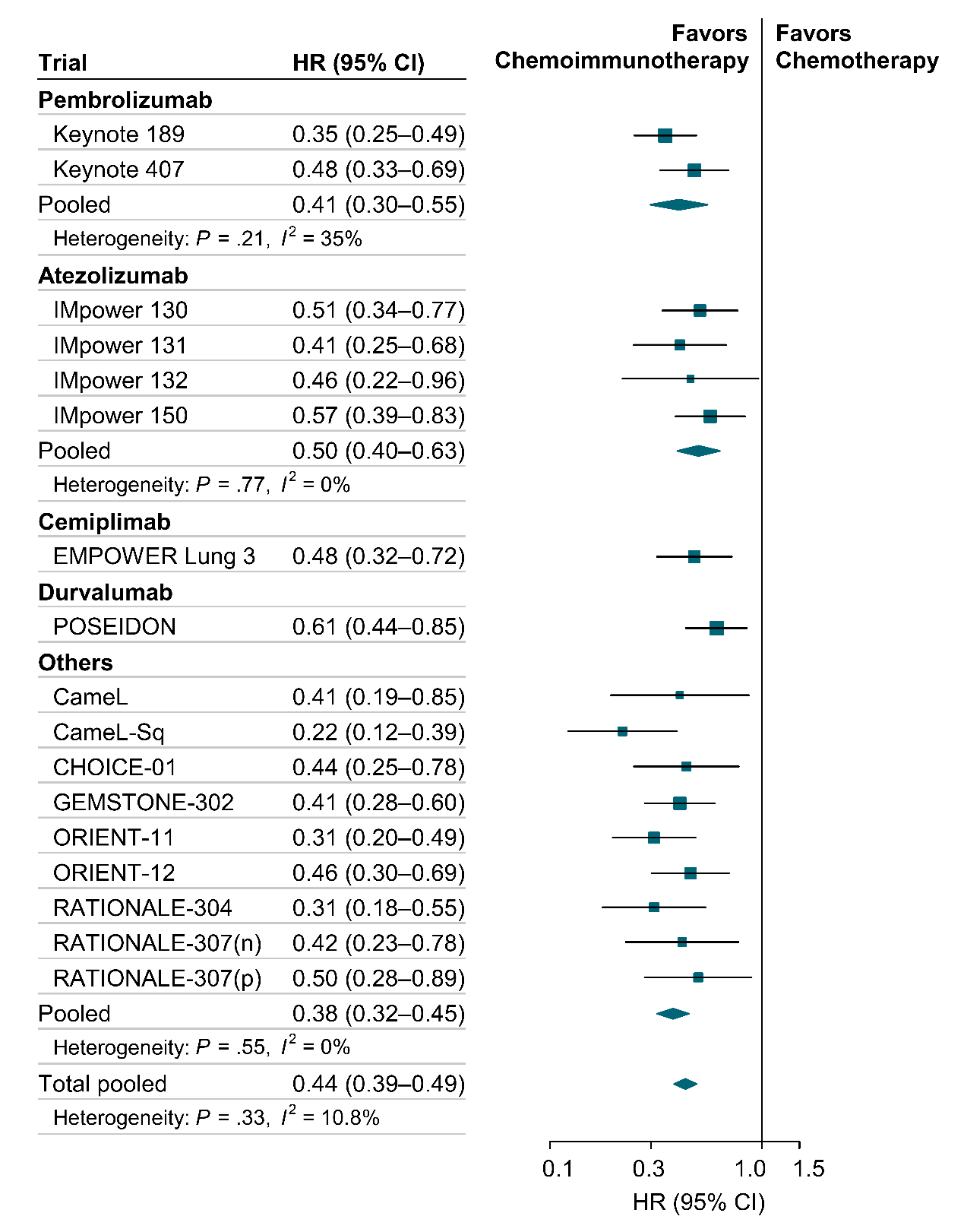
 Abbreviations: CI, confidence interval; HR, hazard ratio; *I*², I-squared statistic (percentage of total variability due to heterogeneity).

**eFigure 4.** Forest Plots of Hazard Ratios for Progression-Free Survival Comparing ICI Monotherapy with Chemotherapy
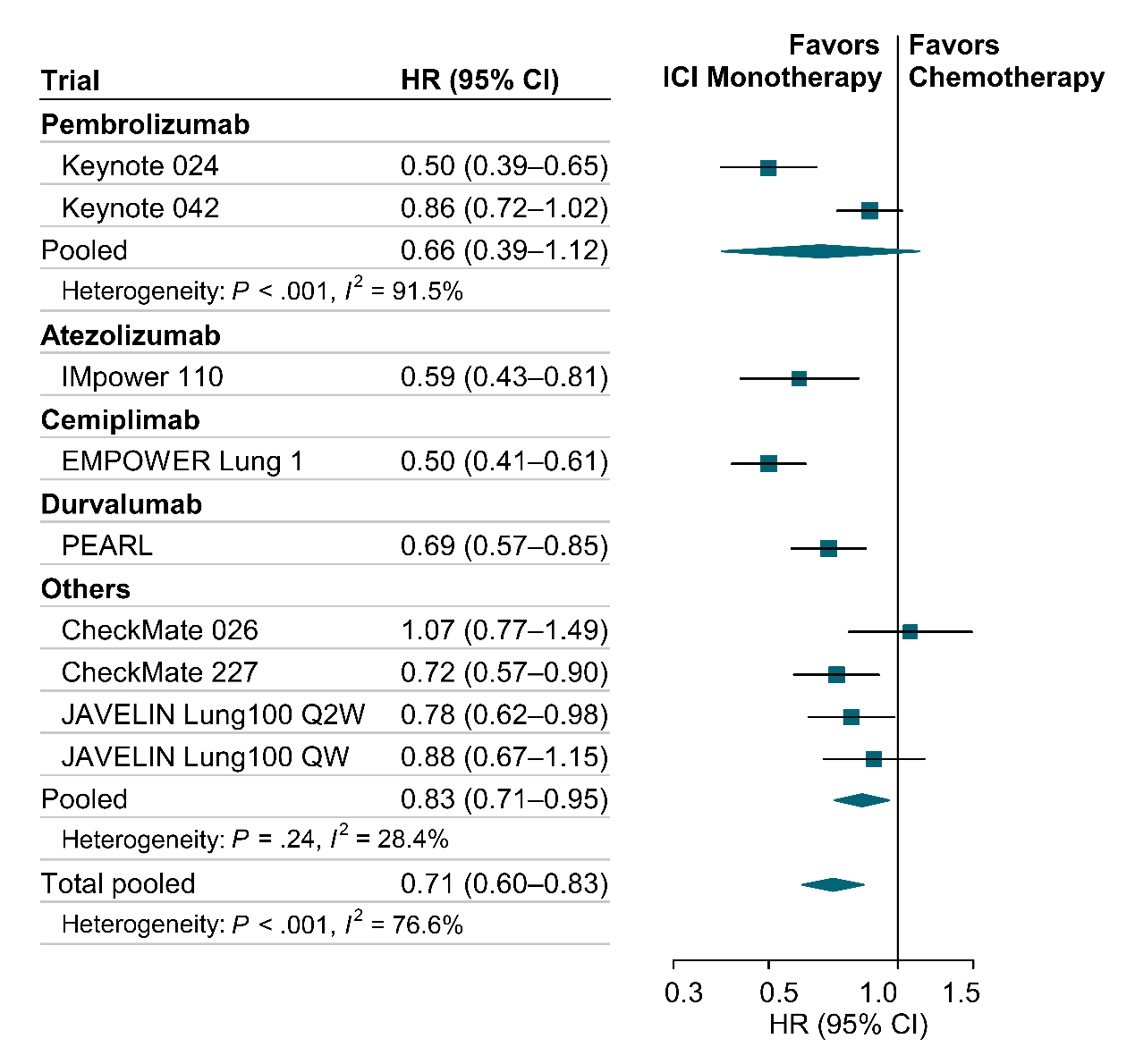
 Abbreviations: CI, confidence interval; HR, hazard ratio; ICI, immune checkpoint inhibitor; *I*², I-squared statistic (percentage of total variability due to heterogeneity).
